# Oncogenic human papillomavirus and anal microbiota in men who have sex with men and are living with HIV in Northern Taiwan

**DOI:** 10.1101/2024.05.08.24307044

**Authors:** Shu-Hsing Cheng, Yu-Chen Yang, Cheng-Pin Chen, Hui-Ting Hsieh, Yi-Chun Lin, Chien-Yu Cheng, Kuo-Sheng Liao, Fang-Yeh Chu, Yun-Ru Liu

**Affiliations:** Department of Infectious Diseases, Taoyuan General Hospital, Ministry of Health and Welfare, Taoyuan, Taiwan; School of Public Health, Taipei Medical University, Taipei, Taiwan; Joint Biobank, Office of Human Research, Taipei Medical University, Taipei, Taiwan; Institute of Clinical Medicine, National Yang Ming Chiao Tung University, Taipei, Taiwan; Graduate Institute of Clinical Medicine, College of Medicine, Taipei Medical University, Taipei, Taiwan; Institute of Public Health, National Yang Ming Chiao Tung University, Taipei, Taiwan; Department of Pathology, Taoyuan General Hospital, Ministry of Health and Welfare, Taoyuan, Taiwan; Department of Clinical Pathology, Far Eastern Memorial Hospital, New Taipei City, Taiwan; Graduate School of Biotechnology and Bioengineering, Yuan Ze University, Taoyuan City, Taiwan; School of Medical Laboratory Science and Biotechnology, Taipei Medical University, Taipei, Taiwan

## Abstract

Few studies have demonstrated the interplay between human immunodeficiency virus (HIV), anal human papillomavirus (HPV), and anal microbiota, especially in persons living with HIV who are men who have sex with men. We therefore explored these interrelationships in a cohort of persons living with HIV, mainly comprising men who have sex with men. HPV genotyping using a commercial genotyping kit and ThinPrep cytology interpreted by Bethesda systems were performed on samples from 291 patients. Samples were characterized by high-throughput sequencing of dual-index barcoded 16s rRNA (V3–4). Bacterial diversity was diminished in individuals living with HIV with CD4+ T cells <500 cells/µL and anal cytology yielding atypical squamous cells of undetermined significance or higher grades (ASCUS+) with detectable HPV 16/18 compared with those with CD4+ T cells ≥500 cells/µL with ASCUS+ and HPV 16/18 and those with normal anal cytology or inflammation without HPV 16/18. Enterobacteriaceae, Ruminococcus, and Bacilli were significantly abundant in persons living with HIV with CD4+ T cells <500 cells/µL with ASCUS+ and HPV 16/18. Bacterial diversity, composition, and homogeneity of dispersion were different in individuals living with HIV with low CD4+ T cells with ASCUS+ and HPV 16/18. These patients should be followed up for the development of anal precancerous lesions.

## Introduction

Human papillomavirus (HPV) is associated with cancer, including cervical, head and neck, and genital cancers, and anal squamous cell carcinoma [1,2]. Certain populations have been noted to have a higher risk of developing anal cancer, namely persons living with human immunodeficiency virus (HIV) and men who have sex with men (MSM) [3]. Persistent oncogenic HPV infections were noted in persons living with HIV (PLWH), especially with immunocompromised status (CD4 + T cell counts less than 200 µL/mL) [4]. Furthermore, the incidence of HPV-related cancer continues to increase in the era of highly active antiretroviral therapy (HAART) in PLWH [2,5].

Recently, there have been many studies on the symbiotic relationship between the host and indigenous microbiota in establishing viral infections and the progression of virus-related diseases. Changes in commensal microbiota may affect the prognosis of influenza, norovirus, and COVID-19 [6–8]. Dysbiosis has been implicated in carcinogenic pathways, such as chronic B or C hepatitis, hepatocellular carcinoma [9–11], HPV, and cervical carcinogenesis in women [12,13]. Intestinal dysbiosis and chronic immune activation interact in PLWH [14,15]. *Peptostreptococcus* and Fusobacteria have been implicated in the pathogenesis of colorectal cancers [16,17] and are associated with poor prognosis [18].

There are fruitful studies on the interaction between the microbiota, HPV acquisition and persistence, and the development of cervical cancer [13,19,20]. Women with cervical intraepithelial neoplasia exhibit a microbiota profile with increased bacterial diversity and a reduced abundance of *Lactobacillus* spp. However, few studies have demonstrated the interplay between HIV, anal HPV, and anal microbiota, especially in PLWH who are MSM [20–22]. We explored the interrelationships between HIV, anal HPV, anal precancerous lesions, and anal microbiota in a PLWH cohort comprising mainly of MSM.

## Materials and Methods

### Study participants

Between Jun 1^st^, 2018 and Dec 31^st^, 2021, PLWH who were MSM and visited one of the outpatient clinics of Taoyuan General Hospital, Taiwan, voluntarily enrolled in this study. Taoyuan General Hospital is a 1,020-bed referral hospital in northern Taiwan serving 2.3 million people, more than 4,300 of whom live with HIV [23]. After providing written informed consent, the participants completed a self-administered questionnaire that addressed their education level; marital status; sexual behavior (heterosexuality or homosexuality, lifetime number of sexual partners, number of new sexual partners within the previous 6 months, frequency of receptive anal sex [always, often, occasionally, seldom, or never], frequency of condom use during anal sex [always, often, occasionally, seldom, or never], frequency of chemsex [always, often, occasionally, seldom, or never]; self-reported sexually transmitted infections (STIs) within the previous 6 months (syphilis, gonorrhea, chlamydial urethritis, condyloma acuminata, amebic colitis/liver abscess, or other clinical diagnoses of STIs), and history of HPV vaccination. Saline-wetted Dacron swabs (Amplicor STD Swab Specimen Collection and Transport Set; Roche Molecular Systems, Branchburg, NJ, USA) were inserted approximately 5 cm above the anal verge. Rectal swabs were immediately rinsed in vials of PreservCyt solution (Hologic Inc., Marlborough, MA, USA) and Omnigene collection tubes (OMR-130; DNA Genotek Inc., Stittsville, ON, Canada). This study was approved by the Institutional Review Board of Taoyuan General Hospital (IRB#: TYGH106042, TYGH10743 and TYGH108050).

### Anal Pap smears

Anal cytology samples were prepared using thin-preparation Pap smears (ThinPrep; Hologic Inc., Marlborough, MA, USA) and sent to a certified laboratory for blinded interpretation by two cytopathology technicians and two pathologists. The results were classified according to the 2001 Bethesda System [24]. We considered the following as anal cellular dysplasia: atypical squamous cells of undetermined significance (ASCUS), low-grade squamous intraepithelial lesions (LSILs), HSILs, and atypical squamous cells that could not exclude HSIL (ASC-H). The cells were preserved in PreservCyt solution and stored at −70°C for DNA testing.

### Human papillomavirus genotyping

HPV genotyping was performed using polymerase chain reaction (PCR) (Cobas HPV; Roche Molecular Systems, Branchburg, NJ, USA). Fourteen types of HPV been detected, including oncogenic types 16 and 18 (as well as types 31, 33, 35, 39, 45, 51, 52, 56, 58, 59, 66, and 68).

### Targeted amplicon library preparation and sequencing

The 16S rRNA gene amplification and library construction were performed according to Illumina’s recommended protocols [25]. Briefly, the universal primers 341F (5′-CCTACGGGNGGCWGCAG-3′) and 805R (5′-GACTACHVGGGTATCTAATCC-3′) containing Illumina overhang adapter sequences in the forward (5′-TCGTCGGCAGCGTCAGATGTGTATAAGAGACAG-3′) and reverse (5′-GTCTCGTGGGCTCGGAGATGTGTATAAGAGACAG-3′) primers were used to amplify the V3–V4 region of bacterial 16S rRNA using a limited cycle PCR. A Nextera XT Index kit (Illumina Inc., San Diego, CA, USA) was used to add Illumina sequencing adapters and dual-index barcodes to the amplicon targets. Quantification and quality of the sequencing libraries were checked using a QSep100 Analyzer (BiOptic Inc., New Taipei city, Taiwan). Finally, the libraries were normalized and pooled in an equimolar ratio and sequenced using an Illumina Miseq instrument, generating paired-end reads of 300 bases in length.

### 16S rRNA gene sequence analysis

Universal primer sequences and low-quality reads were trimmed using Cutadapt (v1.18) [26] and then processed and analyzed using the DADA2/phyloseq workflow in the R environment. Briefly, filtering, trimming, dereplication, and denoising of forward and reverse reads were performed using DADA2 (v1.10.0) [27]. Processed reads were merged, and chimeras were subsequently removed from the cleaned full-length amplicons. Taxonomic assignment of the inferred amplicon sequence variants (ASVs) was performed using the SILVA reference database (v138) [28] with a minimum bootstrap confidence of 80. Multiple sequence alignments of ASVs were performed using DECIPHER (v2.8.1) [29], and a phylogenetic tree was constructed using RaxML (v8.2.11) [30]. The frequency table, taxonomy, and phylogenetic tree information were used to create a phyloseq object for downstream bacterial community analyses using phyloseq (v1.26.0) [31]. The R package Generalized Unique Fraction (GUniFrac) was used to calculate different versions of Unique Fraction (UniFrac) distances. Microbiota enrichment analysis was conducted using the Linear Discriminant Analysis (LDA) effect size (LefSe) method and visualized as a cladogram using GraPhlAn.

### Statistical analyses

Demographic data are presented as mean ± standard deviation (SD) for continuous variables and percentiles for discrete variables. The distribution of the cytology grading was calculated, and the HPV genotype results were analyzed. Chi-square tests were used to compare categorical variables and Student’s *t*-tests were used to compare continuous variables. Anal samples were divided into six groups; group 1: samples from PLWH whose CD4+ T cell counts were < 500 cells/µL with anal cytology yielding ASCUS or higher grades and detectable HPV 16/18; group 2: samples from PLWH whose CD4+ T cell counts were ≥ 500 cells/µL with anal cytology yielding ASCUS or higher grades with detectable HPV 16/18; group 3: samples from PLWH who had anal cytology yielding ASCUS or higher grades without HPV 16/18 detection, group 4: samples from PLWH whose CD4+ T cell counts were < 500 cells/µL and who had normal anal cytology or inflammation with detectable HPV 16/18; group 5: samples from PLWH whose CD4+ T cell counts ≥ 500 cells/µL and who had normal anal cytology or inflammation with detectable HPV 16/18; and group 6: samples from PLWH who had normal anal cytology or inflammation without HPV 16/18 detection. P < 0.05 was considered statistically significant. All statistical analyses were conducted using SAS 9.3 software (SAS Institute, Inc., Cary, NC, USA).

## Results

In total, 338 anal samples were collected. However, 38 (11.2%) failed cytology interpretation or HPV genotyping tests, and 9 (2.7%) were from heterosexual PLWH. Finally, 291 (86.1%) samples collected from PLWH who were MSM were analyzed (Table 1).

**Table 1.** Demographic characteristics, behavior risks, and immunologic factors for the study cohort of men who have sex with men and live with HIV in northern Taiwan.

| Characteristics | Patients<br>N=291 | Percentage<br>or SD |
| --- | --- | --- |
| Age, mean (SD) | 39.26 | (10.06) |
| Educational year |  |  |
| 6 to 9 | 14 | 4.8% |
| >9 to 12 | 80 | 27.5% |
| >12 | 197 | 67.7% |
| Marriage |  |  |
| single | 271 | 93.1% |
| others | 20 | 6.9% |
| HIV years, mean (SD) | 8.55 | (5.55) |
| Smoking | 115 | 39.5% |
| Drinking | 116 | 39.9% |
| Betelnut | 9 | 3.1% |
| Lifetime sexual partner, mean (SD) | 21.62 | (50.74) |
| Sexual partner in 1/2 y, mean (SD) | 2.17 | (4.33) |
| Condom use during anal sex |  |  |
| Every time, often | 156 | 53.6% |
| Occasional | 114 | 39.2% |
| Rare, never | 21 | 7.2% |
| Met sexual partners on web | 196 | 67.4% |
| STDs in 1/2 y | 85 | 29.2% |
| Circumcision | 52 | 17.9% |
| HPV vaccination | 62 | 21.3% |
| Substance use in 1/2 y | 42 | 14.4% |
| Ever participated in chemsex | 95 | 32.6% |
| Current CD4+ T cell count, mean (SD) | 607.62 | (267.11) |
| Undetectable viral load (VL<50) | 267 | 91.8% |

Among the anal samples from the 291 participants, 199 (68.4%) had detectable oncogenic HPVs, 92 (31.6%) did not, and 68 (23.4%) were positive for HPV type 16/18.

Among the anal samples from the 291 participants, 133 (45.7%) yielded cytology interpretations with ASCUS or higher grades and 158 (52.3%) with normal anal cytology or inflammation.

Among the 133 samples with ASCUS or higher grades in cytology findings, 23 (17.3%) did not have oncogenic HPVs, but 110 (82.7%) had detectable oncogenic HPVs. In the 110 samples with oncogenic HPVs and ASCUS or higher grades, 66 (60%) were from PLWH with CD4 +T cell counts of ≥ 500 cells/µL, 44 (40%) had CD4+ T cell counts of < 500 cells/µL; and 46 (41.8%) had detectable HPV 16/18. In the 46 samples with HPV 16/18 and ASCUS or higher grades, 25 (54.3%) were from PLWH with CD4+ T cell counts of ≥ 500 cells/µL (group 2) and 21 (45.7%) were from PLWH with CD4+ T cell counts of <500 cells/µL (group 1). Anal samples with ASCUS or higher grade and without HPV 16/18 were in group 3.

Of the 158 anal samples with normal or inflamed cytology, 69 (43.7%) did not have oncogenic HPVs, but 89 (56.3%) had detectable oncogenic HPVs. Of the 89 samples with oncogenic HPVs and normal cytology findings or inflammation, 54 (60.7%) were from PLWH with CD4+ cell counts of ≥ 500 cells/µL, 35 (39.3%) were from PLWH with CD4 + T cell counts of <500 cells/µL; and 22 (24.7%) had detectable HPV 16/18. Of the 22 samples with HPV 16/18 and normal cytology findings or inflammation, 9 (40.9%) were from PLWH with CD4+ T cell counts of ≥ 500 cells/µL (group 5) and 13 (59.1%) were from PLWH with CD4+ T cell counts of <500 cells/µL (group 4). Anal samples with normal or inflammation cytology and without HPV 16/18 were included in group 6.

We did not find a clear difference in alpha diversity metrics, which implicated the richness and evenness of bacterial taxa within a community, measured by observed, chao index, Shannon, and Simpson methods, between samples from PLWH who have anal cytology yielding ASCUS or higher grades with detectable HPV 16/18 and CD4+ T cell counts of < 500 cells/µL (group 1) and samples from other groups (observed p = 0.038 for group 1 vs group 2; p = 0.014 for group 1 vs group 6; Chao1 p = 0.041 for group 1 vs group 6; Fig 1).

**Fig 1.**
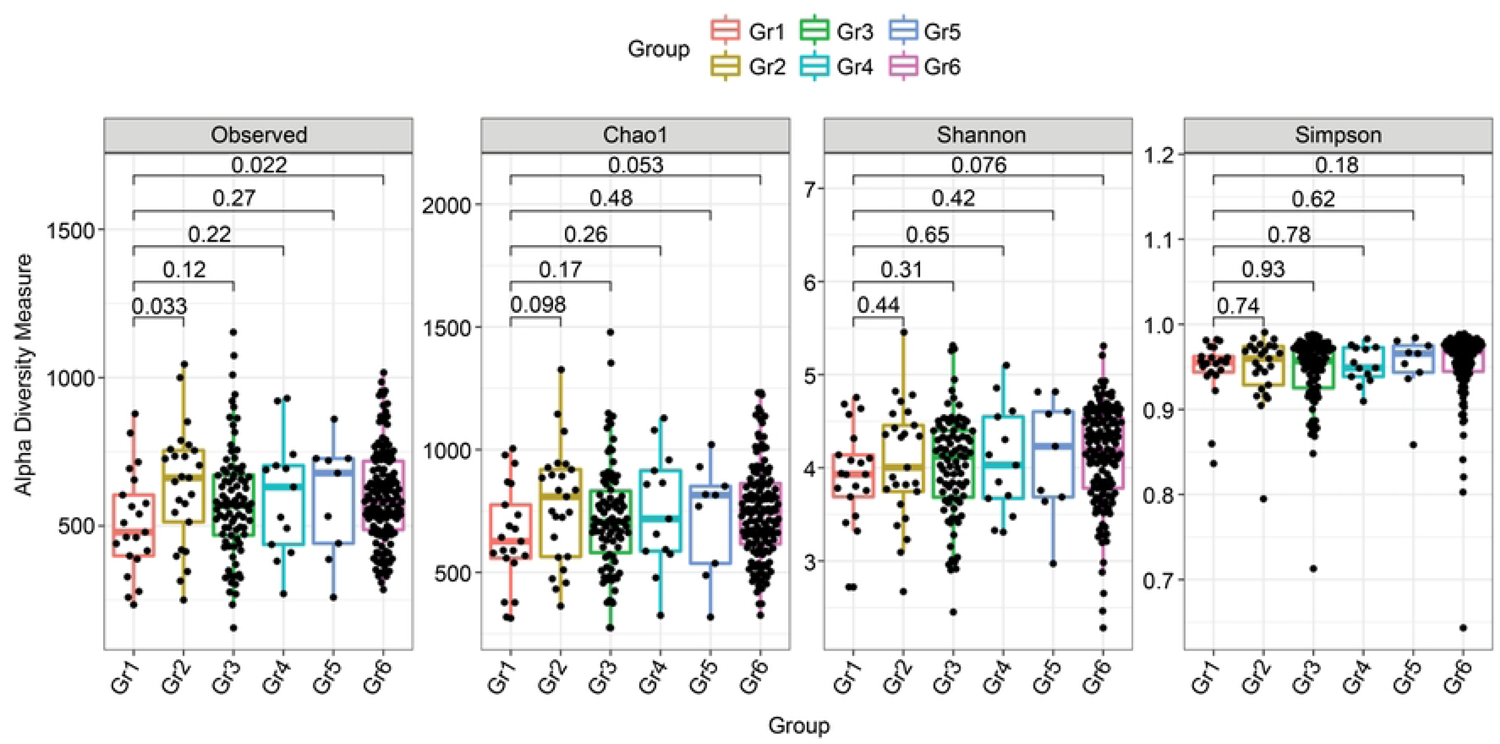
Alpha diversity comparing group 1 and groups 2, 3, 4, 5, and 6. Observed p = 0.038 for group 1 vs group 2, p = 0.014 for group 1 vs group 6; Chao1 p = 0.041 for group 1 vs group 6. Note: group 1: PLWH whose CD4+ T cell counts were < 500 cells/µL with anal cytology yielding ASCUS or higher grades and detectable HPV 16/18; group 2: PLWH whose CD4+ T cell counts were ≥ 500 cells/µL with anal cytology yielding ASCUS or higher grades with detectable HPV 16/18; group 3: PLWH who had anal cytology yielding ASCUS or higher grades without HPV 16/18 detection, group 4: PLWH whose CD4+ T cell counts were < 500 cells/µL and who had normal anal cytology or inflammation with detectable HPV 16/18; group 5: PLWH whose CD4+ T cell counts were ≥ 500 cells/µL and who had normal anal cytology or inflammation with detectable HPV 16/18; and group 6: PLWH who had normal anal cytology or inflammation without HPV 16/18 detection. ASCUS, atypical squamous cells of undetermined significance; Gr, group; HPV, human papillomavirus; PLWH, persons living with HIV.

We also examined beta diversity, which reflects the extent of variation in microbial community composition between the two groups, using Principal Co-ordinates Analysis (PcoA) on unweighted UniFrac and weighted UniFrac, PcoA on variance-adjusted weighted UniFrac, PcoA on GuniFrac with alpha 0.5, Non-metric Multidimensional Scaling (NMDs) on Bray-Curtis distance (VST), Adonis tests (to examine whether composition between groups was similar), and the Betadisper test (to test the homogeneity of dispersion between groups, which is an assumption of Adonis). We found differences at this level between samples from group 1 and those from the other groups (NMDs on Bray-Curtis distance: Adonis test p = 0.0639 [group 1 vs group 2]; NMDs on Bray-Curtis distance: Adonis test 0.0149 [group 1 vs group 3]; PcoA on unweighted UniFrac: Betadisper test p = 0.038 [group 1 vs group 5]; PcoA on unweighted UniFrac: Adonis p = 0.014 [group 1 vs group 6]; PcoA on variance adjusted weighted UniFrac: Adonis p = 0.017 [group 1 vs group 6]; PcoA on GuniFrac with alpha 0.5: Adonis p = 0.030 [group 1 vs group 6]; and NMDs on Bray-Curtis distance (VST): Adonis tests p = 0.0006 [group 1 vs group 6]). For the six groups, plots of NMDs on the Bray-Curtis distance (VST) showed significant composition differences, as tested by the Adonis test (p = 0.0019; **Fig 2a–d**).

**Fig 2.**
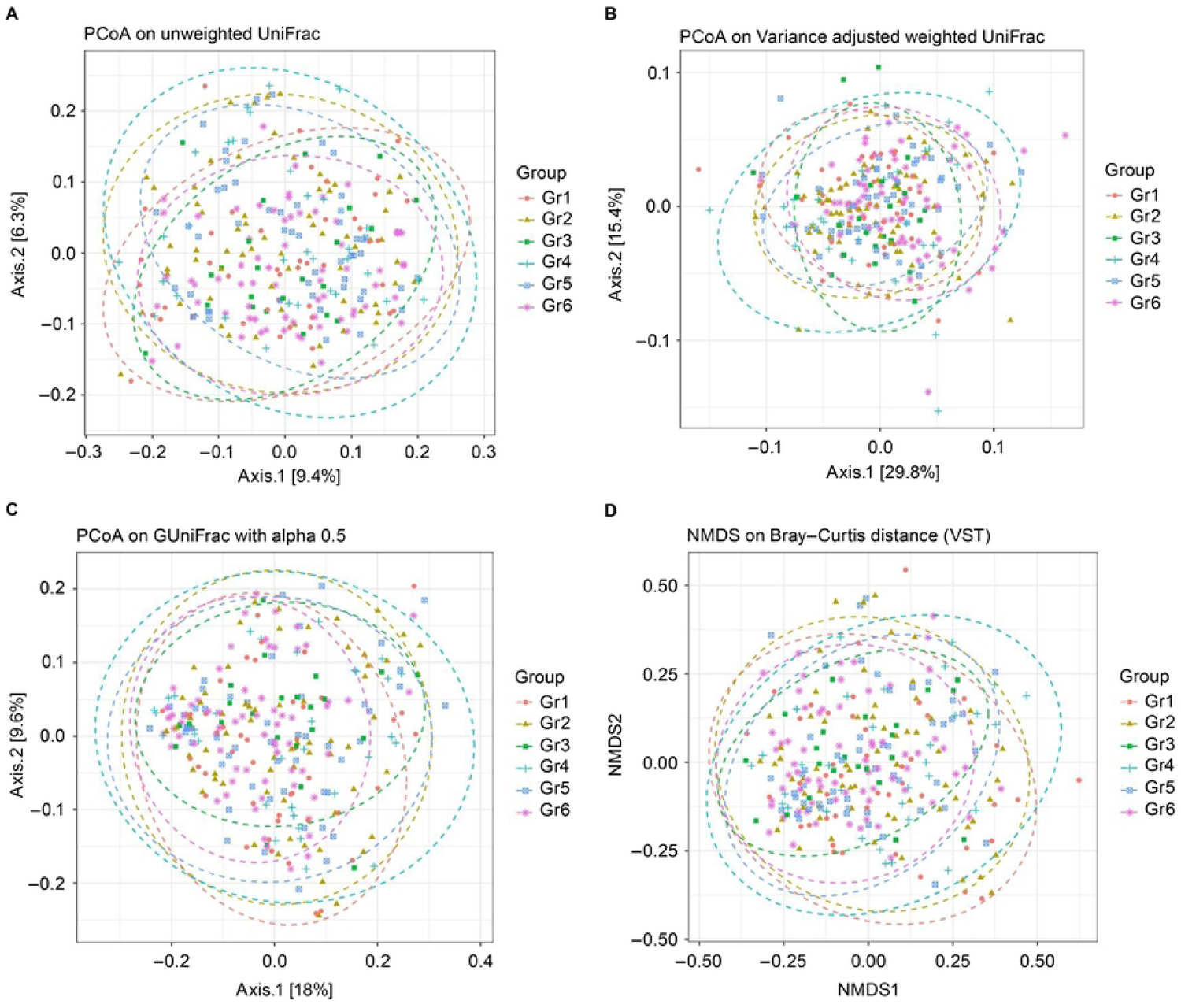
Beta diversity comparing group 1 and groups 2, 3, 4, 5, and 6. 2a: PcoA on unweighted UniFrac: Betadisper test p = 0.038 (group 1 vs group 5), Adonis p = 0.014 (group 1 vs group 6); 2b: no significance; 2b: PcoA on variance adjusted weighted UniFrac: Adonis p = 0.017 (group 1 vs group 6); 2c: PcoA on GuniFrac with alpha 0.5: Adonis p = 0.030 (group 1 vs group 6); and 2d: NMDs on Bray-Curtis distance: Adonis test p = 0.0639 (group 1 vs group 2); Adonis test 0.0149 (group 1 vs group 3); Adonis tests p = 0.0006 (group 1 vs group 6). Note: group 1: PLWH whose CD4+ T cell counts were < 500 cells/µL with anal cytology yielding ASCUS or higher grades and detectable HPV 16/18; group 2: PLWH whose CD4+ T cell counts were ≥ 500 cells/µL with anal cytology yielding ASCUS or higher grades with detectable HPV 16/18; group 3: PLWH who had anal cytology yielding ASCUS or higher grades without HPV 16/18 detection, group 4: PLWH whose CD4+ T cell counts were < 500 cells/µL and who had normal anal cytology or inflammation with detectable HPV 16/18; group 5: PLWH whose CD4+ T cell counts were ≥ 500 cells/µL and who had normal anal cytology or inflammation with detectable HPV 16/18; and group 6: PLWH who had normal anal cytology or inflammation without HPV 16/18 detection. ASCUS, atypical squamous cells of undetermined significance; Gr, group; HPV, human papillomavirus; NMDs, Non-metric Multidimensional Scaling; PcoA: Principal Co-ordinates Analysis; PLWH, persons living with HIV.

We used the LefSe discovery tool to identify the predictors of abnormal anal cytology. This method addresses the issue of identifying microorganisms that consistently explain the differences between ≥2 microbial communities. When comparing groups 1 and 2, LefSe showed abundant Enterobacterales, Enterobacteriaceae, and *Mycoplasma* in group 1 and abundant *Finegoldia* in group 2 (Fig 3a). When comparing groups 1 and 3, LefSe showed abundant Enterobacterales and Enterobacteriaceae in group 1, and abundant *Alloprevotella* and Erysipelatoclostridiaceae in group 3 (Fig 3b). When comparing groups 1 and 4, LefSe showed abundant *Ruminococcus* and *Sutterella* in group 1 and abundant Porphyromonadaceae, *Porphyromonas*, and *Fenollaria* in group 4 (Fig 3c). When comparing groups 1 and 5, LefSe did not show significant findings. When comparing groups 1 and 6, LefSe showed abundant Enterobacterales, Enterobacteriaceae, and Bacilli in group 1, and abundant Bacteroidota, Bacteroidia, Bacteroidales, and Clostridia in group 6 (Fig 3d).

**Fig 3.**
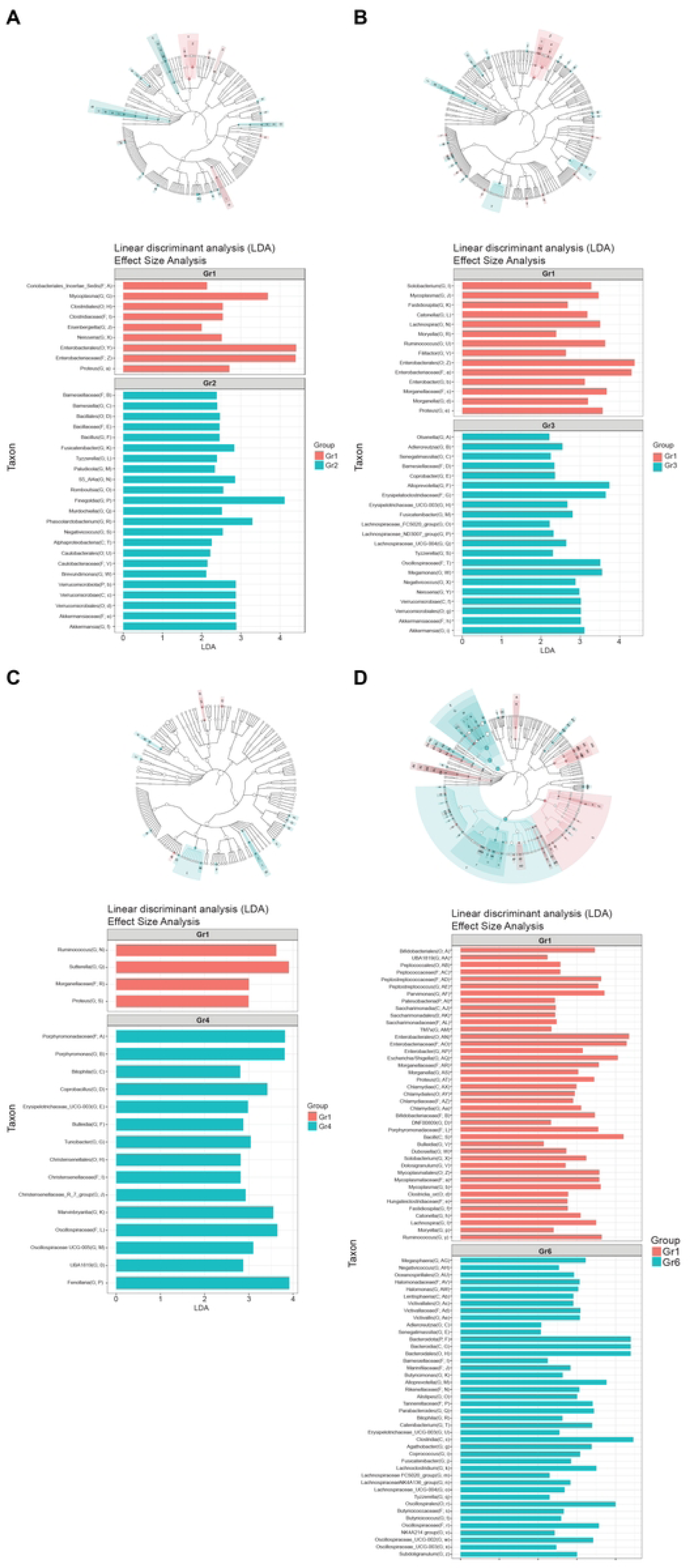
Linear discriminant analysis effect size comparing group 1 and groups 2, 3, 4, 5, and 6 and cladogram. 3a: group 1 vs group 2; 3b: group 1 vs group 3; 3c: group 1 vs group 4, 3d: group 1 vs group 6. Note: group 1: PLWH whose CD4+ T cell counts were < 500 cells/µL with anal cytology yielding ASCUS or higher grades and detectable HPV 16/18; group 2: PLWH whose CD4+ T cell counts were ≥ 500 cells/µL with anal cytology yielding ASCUS or higher grades with detectable HPV 16/18; group 3: PLWH who had anal cytology yielding ASCUS or higher grades without HPV 16/18 detection, group 4: PLWH whose CD4+ T cell counts were < 500 cells/µL and who had normal anal cytology or inflammation with detectable HPV 16/18; group 5: PLWH whose CD4+ T cell counts were ≥ 500 cells/µL and who had normal anal cytology or inflammation with detectable HPV 16/18; and group 6: PLWH who had normal anal cytology or inflammation without HPV 16/18 detection. ASCUS, atypical squamous cells of undetermined significance; Gr, group; HPV, human papillomavirus; PLWH, persons living with HIV.

## Discussion

The interaction between microbiota, HPV acquisition and persistence, and the development of cervical cancer in women has been researched in detail [13,19,20]. Based on the presence or absence of a particular *Lactobacillus* species, Ravel et al. [32] assigned five different community state types (CSTs); CST I, II, III, and V were dominated by *Lactobacillus crispatus*, *L. gasseri*, *L. iners*, and *L. jensenii*, respectively, and CST IV (bacterial vaginosis-associated bacteria) was a heterogeneous group characterized by the depletion of *Lactobacillus* spp. in the presence of anaerobic species such as *Gardnerella*, *Megasphera*, *Sneathia*, and *Prevotella*. The vaginal ecosystem is dominated by *Lactobacillus* spp. Lactobacilli provide broad-spectrum protection by producing lactic acid, bacteriocins, and biosurfactants and by adhering to the mucosa, which forms barriers against pathogenic infections in the vaginal microenvironment [33]. If an imbalance in this defense system occurs, physicochemical changes arise, inducing selective pressure on the microbiota (depletion of certain *Lactobacillus* and increase in richness of microbiota) and subsequent histological alterations of the vaginal mucosa and the cervical epithelium (squamous intraepithelial neoplasm) [34,35]. A meta-analysis including 12 studies showed bacterial vaginosis to be associated with higher rates of HPV infection (odds ratio 1.43, 95% CI 1.11– 1.84) [36], implying that a *Lactobacillus*-depleted microbiome may contribute to HPV persistence in women cervix. Bacterial vaginosis may be associated with the production of reactive oxygen species that may cause double-stranded DNA damage in the host genome and the HPV episome, facilitating HPV integration and ultimately neoplastic transformation, a mechanism also employed by the HPV E6 oncoprotein [34]. Women with cervical intraepithelial neoplasia exhibit a microbiota profile with increased bacterial diversity and a reduced abundance of *Lactobacillus* spp. [13,19]. These findings were similar in women who are PLWH and had cervical lesions [37,38].

In the pathophysiology of infectious diseases, pathogenic colonization and infection involve dynamic interactions between microbes and the microbiome, host, and environmental factors [39]. This biology is also applicable to PLWH who are MSM. A small study of 42 PLWH who were MSM revealed that *Peptostreptococcus*, *Campylobacter*, and *Gardnerella* were associated with anal precancerous lesions [21]. Another study involving 113 Nigerian MSM found that a higher proportion of MSM with prevalent HPV16 was associated with a cluster enriched in Sneathia from the family Fusobacteriaceae [22]. However, there were only six PLWH treated with HAART with viral suppression in the study by Nowak et al. [22]. Another study showed that Ruminococcaceae NK4A214 group, *Alloprevotella* genus, *Prevotella melanonigenica*, and Ruminococcaceae UCG-014 were the most predictive of biopsy-proven high-grade squamous intraepithelial lesions (HSIL) [40].

In the present study, diminished richness of bacterial diversity was noted in PLWH with CD4+ T cell counts of < 500 cells/µL and anal cytology yielding ASCUS or higher grades with detectable HPV 16/18 (group 1) compared with other groups, i.e., PLWH with CD4+ T cell counts of ≥ 500 cells/µL and anal cytology yielding ASCUS or higher grades with detectable HPV 16/18 (group 2), and PLWH who have normal anal cytology or inflammation without detectable HPV 16/18 (group 6). The bacterial composition and homogeneity of the dispersion also differed between the groups. Enterobacteriaceae, Proteus, Morganellaceae, Mycoplasma, Sutterella, Ruminococcus, and Bacilli were significantly abundant in PLWH whose CD4+ T cell counts were < 500 cells/µL and who had anal cytology yielding ASCUS or higher grades with detectable HPV 16/18 (group 1). In contrast, Bacteroidales and Clostridia were abundant in PLWH with anal cytology, yielding normal or inflammatory results without detectable HPV 16/18 (group 6).

HIV infection also frequently correlates with increased tissue and circulating measures of inflammation, such as CD14, IL-6, and CD38+HLA-DR+CD8+ T cells in the gut [41], as well as increases in traditionally pathogenic bacteria such as Enterobacteriaceae, including Morganellaceae and *Proteus* [42,43].

*Mycoplasma* is the smallest intracellular bacterium that establishes persistent intracellular infections that can lead to inflammatory cytokine-mediated tissue injury. In addition, m*ycoplasma* infection allows direct interaction with HPV during co-infection of epithelial cells [44], an observation consistent with our findings. *Sutterella* is a gram-negative, non-spore-forming rod grown under anaerobic or microaerophilic conditions. It belongs to the family Betaproteobacteria, is proinflammatory [45], and could explain our findings regarding abundance of *Sutterella* in the anal samples of group 1 patients.

*Ruminococcus*, a gram-positive, spore-forming, anaerobic rod, is a polyphyletic genus with species belonging to two Firmicutes families, Lachnospiraceae and Ruminococcaceae, capable of degrading cellulose. Our findings are consistent with those of Ron et al., who described *Ruminococcus*-predicting HSIL [40]. In contrast, a decreased abundance of *Ruminococcus* was previously noted in the guts of PLWH [46]. Bacilli have been reported to correlate with the progression of HPV-related cervical lesions [13] and may therefore deserve attention with regard to the progression of anal lesions.

The strength of this study lies in its interpretation of the anal microbial compositions of a large cohort of PLWH who are MSM and had different stages of HPV-related disease using high-throughput sequencing. The limitations of this study include its cross-sectional design. Hence, we could not conclude a causal relationship between the anal microbiota and HPV infection or AIN. Second, nearly 100% of the study cohort received HAART; therefore, the findings of this study cannot be generalized to PLWH without HAART. Third, previous studies have mentioned the contribution of Fusobacterium and Sneathia to HPV-related lesions [22,39], which could not be proven in this study. Ethnic or environmental factors may contribute to these findings [13,22]. Finally, the participants in this study were from a single urban tertiary care hospital, and the results cannot be generalized to different populations.

In conclusion, the diminished richness, composition, and homogeneity of dispersion of bacterial diversity were different in PLWH with low CD4+ T cell counts (< 500 cells/µL) and who have anal cytology yielding ASCUS or higher grades with detectable HPV 16/18. These patients should be followed-up for the development of anal precancerous lesions. Furthermore, the anal swab signature microbiota profiles, such as *Lactobacillus* in the cervix, warrant further exploration.

## Data Availability

The data that support the findings of this study are available in the Figshare database, doi: 10.6084/m9.figshare.25375087.

https://doi.org/10.6084/m9.figshare.25375087

## Acknowledgements

We acknowledge the Core Instrument Center of the Taipei Medical University for providing the next generation sequence service. We also thank the patients and the care team of Comprehensive AIDS Care Center at Taoyuan General Hospital, Ministry of Health and Welfare.

